# Cardiac and cardiometabolic multimorbidity and risk of all-cause dementia: a nested case-control study using routine primary care data (N=347,436)

**DOI:** 10.64898/2026.11.26.26369039

**Authors:** Thomas Donaldson, Jordan Canning, Laura M. Lyall, Frederick K. Ho, Terry Quinn, Donald M. Lyall

**Affiliations:** School of Health and Wellbeing, University of Glasgow, Glasgow, UK; School of Cardiovascular and Metabolic Sciences, University of Glasgow, Glasgow, UK

**Keywords:** dementia, cardiovascular disease, multimorbidity, electronic health records, cardiac, CPRD, epidemiology

## Abstract

**Background.:** Cardiac and cardiometabolic conditions are known risk factors for dementia, however the impact of multimorbidity is not understood. This study assessed the association between individual conditions and subsequent dementia, and tested for synergistic risk conferral from multimorbidity.

**Methods and findings.:** A nested retrospective case-control study was conducted using electronic health record (EHR) data from the Clinical Practice Research Datalink (CPRD). Adults aged ≥60 years with incident all-cause dementia (2012-2020) were matched 1:5 to controls by age, sex, and GP practice to proxy deprivation (57,906 cases; 289,530 controls). Cox proportional hazards regression estimated hazard ratios (HRs) for individual and grouped cardiac and cardiometabolic conditions, with interaction terms testing synergistic risk. Any cardiac condition was associated with 13% increased all-cause dementia risk (HR 1.13, 95% CI 1.10-1.16), and any cardiometabolic condition with a 12% increased risk (HR 1.12, 95% CI 1.10-1.14). Co-occurring diagnoses conferred 22% increased risk (HR 1.22, 95% CI 1.17-1.26). The interaction was sub-additive (HR 0.92, 95% CI 0.87-0.98, P=0.008; RERI interaction coefficient - 0.08); i.e. greatest risk was conferred by a first cardiac/cardiovascular diagnosis. Several individual conditions showed duration-dependent association, e.g. diabetes rose from HR 1.05 (95% CI 1.01-1.09) at 5-10 years, to HR 1.56 (95% CI 1.38-1.75) at ≥15 years. The main limitations are absence of genotype and lifestyle data.

**Conclusions.:** In routine anonymised electronic healthcare data that is representative of the general UK population, we show that most dementia risk is conferred by a first cardiovascular/cardiac diagnosis, with diminishing risk from additional conditions. Increasing risk across time strengthens the case for embedding cognitive surveillance within long-term cardiometabolic disease care.

**Author summary:** *Why was this study done?:* Poorer physical health is a known dementia risk factor. Cardiac conditions (e.g. atrial fibrillation), and cardiometabolic conditions (e.g. diabetes), are each associated with increased dementia risk overall, and cardiometabolic multimorbidity has been linked to accelerated cognitive decline and higher dementia incidence. However, previous multimorbidity studies have largely been based on volunteer cohorts with relatively small multimorbid subgroups or short follow-up; we found no study that had formally tested whether concurrent cardiac and cardiometabolic diagnoses confer synergistic dementia risk.

*What did the researchers do and find?:* We show that in 347,436 participants from UK primary care, a significant amount of dementia risk was conferred by a first prior cardiovascular or cardiac diagnosis - but with diminishing incremental risk from additional conditions. For several conditions, dementia risk escalated with duration, most prominently for diabetes - reaching 56% increased risk at ≥15 years.

*What do these findings mean?:* A first cardiovascular/cardiac diagnosis confers the greatest dementia risk, consistent with overlapping mechanistic pathways largely captured by initial disease. The progressive increase in dementia risk supports embedding cognitive assessment within cardiometabolic care. Limitations include missing data on genetic and lifestyle risk factors, and probable under- recording of dementia in primary care.

## Introduction

Dementia is the collective term for a group of neurological conditions that impact brain function and that are associated with worsening memory, reasoning, and other cognitive abilities over time. The primary risk factor is age, and with populations continuing to grow and age, the currently estimated 50 million people living with dementia globally is projected to increase to over 150 million by 2050 [1]. In 2021, dementia was the seventh leading cause of death globally [2]. In the United Kingdom (UK), the annual cost of dementia - currently £42 billion - is forecast to reach £90 billion by 2040 [3].

With no cure or restorative treatment currently available, the focus is on preventative approaches and action that can minimise risk and delay onset. The latest Lancet Commission on dementia prevention, intervention and care reported that action on 14 modifiable risk factors could prevent almost half of dementia cases [4]. Several of these centre on cardiovascular health: in recent decades research has found that cardiac conditions like heart failure and atrial fibrillation [5,6], and cardiometabolic conditions like diabetes and hypertension [7,8], are independent risk factors for the development of dementia.

This research has enhanced understanding of the ’brain-heart loop’: the bidirectional relationship whereby cardiovascular health impacts cognitive functioning and vice-versa [6,9]. Vascular integrity is essential for cerebral perfusion, and cardiovascular risk factors that impair this integrity can lead to structural and functional brain damage causing cognitive problems [6,9]. Many cardiovascular conditions share risk factors with dementia - including poor diet, physical inactivity, and obesity - and there is evidence that cardiovascular dysfunction promotes neuroinflammation [10].

Despite this established interplay, there has been limited research into the risk associated with cardiovascular multimorbidity (i.e. the presence of two or more long-term conditions). More specifically, the impact of concurrent cardiac and cardiometabolic diagnoses has not been explored. It is possible that risks may be more than additive, i.e. synergistic. However, relatively small sample sizes or short follow-up periods in previous studies have precluded this analysis [11–14].

## Methods

### Study design and data source

Data were from the Clinical Practice Research Datalink (CPRD), a database of longitudinal medical records from primary care practices across the UK. CPRD has data for over 60 million individuals and is broadly representative of the UK population [15]. CPRD has ethical approval from the Health Research Authority (Research Ethics Committee reference 05/MRE04/87) to provide anonymised patient data for observational research, so individual patient consent was not required. Access was approved through the CPRD Research Data Governance process (protocol 20_000212). This study was reported in accordance with the STROBE guidelines and the CODE-EHR minimum dataset framework for studies using healthcare records and registries.

A nested case-control study was conducted. Individuals aged ≥60 years with a diagnosis of dementia were identified and matched to a control group (five controls per case) based on age (year of birth within three years), sex, and GP practice as a proxy for deprivation, using incidence density matching. The study period ran from 1 January 2005 to 31 December 2020, with dementia case ascertainment restricted to 1 January 2012 to 31 December 2020 to ensure a minimum of seven years of prior CPRD registration for all cases. Follow-up for all participants began on 1 January 2005 or the date of practice registration, whichever was later. Exposure diagnoses were ascertained from the full available CPRD record for each participant, extending back to database inception, provided a minimum of five years elapsed between the diagnosis date and the dementia index date. All relevant codes are specified in S1–S6 Tables. Duplications within each of Aurum/Gold, and then across cohorts, were removed.

### Exposures

Cardiometabolic conditions included diabetes (Type 1/Type 2 and associated complications), hyperlipidaemia, and hypertension. Cardiac conditions were broadly defined as coronary heart disease (CHD), angina, atrial fibrillation (AF), coronary artery operations (CAO), heart failure (HF), and myocardial infarction (MI). The categorisation of conditions was based on whether they affect the heart directly (cardiac) or the broader metabolic and circulatory system (cardiometabolic), consistent with prior CPRD dementia research [5]. Exposures were identified using validated CPRD GOLD and Aurum medcodes (S1 and S2 Tables). Exposure diagnoses occurring within five years of the dementia index date were excluded to mitigate reverse causation.

### Outcome

The outcome was all-cause dementia, ascertained using pre-specified validated medcodes identifying a formally recorded diagnosis of dementia or a prescription of dementia medication (e.g. memantine, donepezil; S3 and S4 Tables). All-cause dementia was selected in preference to specific subtypes: there is significant overlap in pathology between subtypes [16], misclassification is common [17], and prior research concludes that all-cause dementia is a valid measure within primary care data [18].

### Statistical analysis

Descriptive analysis of baseline characteristics and exposure prevalence was conducted for cases and controls. Cox proportional hazards models were used to estimate HRs and 95% confidence intervals (CIs) for all-cause dementia associated with each individual condition, with grouped cardiac/cardiometabolic categories, and with a categorical interaction term (levels: cardiac only, cardiometabolic only, both, neither). Follow-up began at the latest of the practice registration date or study start date, and was censored at dementia index date, death, or 31 December 2020. Age, sex, and area-level deprivation (Index of Multiple Deprivation quintile, IMD, based on GP practice postcode) were controlled for by design through the matched case- control selection. Interactions between exposure groups were assessed on the multiplicative scale via a product term in stratified Cox regressions, and on the additive scale based on Relative Excess Risk due to Interaction (RERI) estimates.

The proportional hazards assumption was tested for each exposure using Schoenfeld residuals (P<0.05 defined as a violation). Variables violating the assumption were excluded from grouped category analyses and assessed separately in stratified models across three follow-up periods (5-9.999, 10-14.999, and ≥15 years). We calculated additional cause-specific associations where proportional hazards were violated. Sensitivity analyses were performed after excluding patients with a history of non-dementia neurodegenerative conditions (e.g. Parkinson’s disease, motor neuron disease), demyelinating disorders (multiple sclerosis), and severe mental illness (schizophrenia, bipolar disorder) at any point during the study. As sensitivity analyses, diagnoses recorded before 1 January 1987, which could feasibly be instances of backdating or error, were conservatively excluded as predating the database. All dataset construction was completed in R (version 4.4.2); statistical analysis was completed using Stata/MP 18.0.

## Results

### Study population

Practices that migrated from CPRD GOLD (Vision) to CPRD Aurum, identified from the CPRD VisionToEmisMigrators file, were present in the extract under both their GOLD and Aurum practice identifiers for 159 practices. Because individuals cannot be linked across the two databases, the entire Aurum-side record for each such practice was removed and the GOLD- side retained, excluding 6,318 participants. The combined CPRD Aurum and GOLD dataset included 57,906 eligible dementia cases matched to 289,530 controls (total N=347,436). Most cases were female (34,874 [60.2%]), and the mean age at dementia index was 80.8 years (SD 8.50). The sample skewed slightly more affluent than the general population: 44.0% of participants came from the two least deprived IMD quintiles compared with 35.3% from the two most deprived. Around half of the sample (49.9%) came from GP practices in England; the remainder were from Scotland (27.2%), Wales (17.6%), and Northern Ireland (5.3%). There were 120,098 deaths during the study period (mean age at death 81.8 years, SD 7.7). The median follow-up period was 14.91 years (IQR 10.95-15.92 years).

The most prevalent exposure conditions amongst dementia cases were hypertension (32,670 [56.4%]), diabetes (11,868 [20.5%]), and AF (10,733 [18.5%]). All conditions were more prevalent amongst cases than controls, with the exception of HF (cases 5,144 [8.9%] vs. controls 26,704 [9.2%]). There were significant differences between all exposure conditions in terms of sex, age group, and IMD quintile (all P<0.001). The prevalence of any cardiac diagnosis was greater in men (38.8%) than women (30.2%), while cardiometabolic conditions were more prevalent amongst women (60.1%) than men (57.0%). Descriptive statistics are presented in Table 1. A total of 55,427 exposure diagnoses were excluded from the primary analysis as there were fewer than five years between diagnosis and dementia index.

**Table 1.** Descriptive statistics for cases and controls.

| **Variable** | **Cases, n (%)<br>(n=57,906)** | **Controls, n (%)<br>(n=289,530)** |
|---|---|---|
| Female | 34,874 (60.2) | 174,370 (60.2) |
| Age at index date, mean (SD), years | 80.8 (8.50) | 80.6 (8.60) |
| Hypertension | 32,670 (56.4) | 145,963 (50.4) |
| Diabetes | 11,868 (20.5) | 49,140 (17.0) |
| Atrial fibrillation | 10,733 (18.5) | 42,246 (14.6) |
| Coronary heart disease | 6,375 (11.0) | 27,895 (9.6) |
| Hyperlipidaemia | 6,162 (10.6) | 24,464 (8.4) |
| Myocardial infarction | 5,404 (9.3) | 25,493 (8.8) |
| Heart failure | 5,144 (8.9) | 26,704 (9.2) |
| Coronary artery operations | 4,149 (7.2) | 19,560 (6.8) |
| Angina | 1,867 (3.2) | 7,369 (2.5) |
| Any cardiometabolic condition | 37,742 (65.2) | 166,893 (57.6) |
| Any cardiac condition | 21,831 (37.7) | 94,936 (32.8) |

### Primary analysis - individual conditions

Four of the nine individual exposures met the proportional hazards assumption and were significantly associated with an increased risk of all-cause dementia (Table 2). The greatest risk was conferred by CHD (HR 1.25, 95% CI 1.20-1.29). Hypertension, the most prevalent condition (present in 178,633 [51.4%] of the total sample), was associated with a 17% increased dementia risk (HR 1.17, 95% CI 1.15-1.20). Hyperlipidaemia was associated with a 17% increased risk (HR 1.17, 95% CI 1.13-1.21), and angina with a 17% increased risk (HR 1.17, 95% CI 1.10-1.25). MI did not reach statistical significance (HR 1.03, 95% CI 0.99-1.07, P=0.15). Diabetes, AF, CAO, and HF violated the proportional hazards assumption; their overall HRs are shown in Table 2 for reference. Cause-specific associations are shown in S7 Table; each was associated with increased risk of death, suggesting that apparently protective associations between AF, CAO, and HF with dementia, were due to competing risk of death.

**Table 2.** Hazard ratios (HRs) and 95% confidence intervals for individual exposure conditions and all-cause dementia.

| **Condition** | **HR** | **95% CI** | **P-value** |
|---|---|---|---|
| *Meets proportional hazards assumption* |  |  |  |
| Coronary heart disease | 1.25 | 1.20-1.29 | <0.001 |
| Hypertension | 1.17 | 1.15-1.20 | <0.001 |
| Angina | 1.17 | 1.10-1.25 | <0.001 |
| Hyperlipidaemia | 1.17 | 1.13-1.21 | <0.001 |
| Myocardial infarction | 1.03 | 0.99-1.07 | 0.15 |
| *Violates proportional hazards assumption\** |  |  |  |
| Diabetes | 1.17 | 1.14-1.20 | <0.001 |
| Atrial fibrillation | 0.90 | 0.87-0.93 | <0.001 |
| Coronary artery operations | 0.87 | 0.83-0.91 | <0.001 |
| Heart failure | 0.77 | 0.73-0.81 | <0.001 |

*\*Overall HR shown for reference only; time-stratified results in Table 4.*

### Multimorbidity and interaction analysis

Following grouping of conditions - which met proportional hazards assumptions - into categories (Table 3), individuals with any cardiac condition were at 13% increased risk of all-cause dementia (HR 1.13, 95% CI 1.10-1.16, P<0.001), while individuals with a cardiometabolic diagnosis were 12% more likely to develop dementia compared to those with no conditions (HR 1.12, 95% CI 1.10-1.14, P<0.001). Patients who had both a cardiac and a cardiometabolic diagnosis were at a 22% increased risk (HR 1.22, 95% CI 1.17-1.26, P<0.001) compared to those with no exposure diagnoses - a greater effect than either individual category, but a less- than-additive increase. Individual condition-by-condition interactions are shown in S8 and S9 Tables and show similar results (i.e. non-significant, interaction estimates below 1, and/or negative RERI estimates).

**Table 3.** Hazard ratios for grouped condition categories, their combined effect, and the interaction term.

| **Category (reference: no condition)** | **HR** | **95% CI** | **P-value** |
|---|---|---|---|
| Cardiac only | 1.13 | 1.10-1.16 | <0.001 |
| Cardiometabolic only | 1.12 | 1.10-1.14 | <0.001 |
| Both cardiac and cardiometabolic | 1.22 | 1.17-1.26 | <0.001 |
| Interaction term (cardiac x cardiometabolic) | 0.92 | 0.87-0.98 | 0.008 |

The interaction test showed an HR of 0.92 (95% CI 0.87-0.98, P=0.008) and a RERI estimate of −0.08 (95% CI −0.15 - -0.01, P =0.026), indicating that the interaction effect was antagonistic rather than additive. In other words, there is greater impact from developing a first condition (from a baseline of zero), than from developing a second condition (from a baseline of one). The greatest dementia risk is conferred by a first exposure (cardiac; cardiovascular) diagnosis, with diminishing incremental risk from additional conditions.

### Time-stratified analyses

For each condition, duration-stratified analyses were run by 5-9.999; 10-14.999 and ≥15 years follow-up (Table 4). AF, CAO, and HF were initially associated with a decreased risk of dementia at 5-10 years follow-up (HRs 0.80, 0.74, and 0.66 respectively; all P<0.001). Effects attenuated toward null at 10-15 years (for AF; CAO), with directional increases at ≥15 years that did not reach statistical significance. This pattern is consistent with competing mortality masking a longer-term signal. Other conditions showed monotonic increases across time bands, where risk increased over time for diabetes, CHD, hypertension, and hyperlipidaemia. This was most pronounced for diabetes. While associated with only a 5% increased dementia risk at 5-10 years of follow-up (HR 1.05, 95% CI 1.01-1.09, P=0.021), the hazard increased substantially with longer follow-up: HR 1.25 (95% CI 1.20-1.30, P<0.001) at 10-15 years, and HR 1.56 (95% CI 1.38-1.75, P<0.001) at ≥15 years. These duration-dependent increases were robust across sensitivity analyses.

**Table 4.** Time-stratified hazard ratios for conditions violating the proportional hazards assumption.

|  | 5-9.999 years |  |  | 10-14.999 years |  |  | ≥15 years |  |  |
|---|---|---|---|---|---|---|---|---|---|
| **Condition** | **HR** | **95% CI** | **P** | **HR** | **95% CI** | **P** | **HR** | **95% CI** | **P** |
| Coronary heart disease | 1.20 | 1.14-1.26 | <0.001 | 1.29 | 1.23-1.36 | <0.001 | 1.34 | 1.13-1.59 | <0.001 |
| Heart failure | 0.66 | 0.62-0.71 | <0.001 | 0.88 | 0.82-0.95 | <0.001 | 1.13 | 0.88-1.45 | 0.32 |
| Atrial fibrillation | 0.80 | 0.76-0.84 | <0.001 | 1.00 | 0.95-1.05 | 0.985 | 1.16 | 0.98-1.37 | 0.089 |
| Angina | 1.08 | 0.98-1.18 | 0.11 | 1.27 | 1.15-1.39 | <0.001 | 1.38 | 0.99-1.92 | 0.056 |
| Myocardial infarction | 0.98 | 0.93-1.04 | 0.53 | 1.08 | 1.02-1.15 | 0.007 | 1.04 | 0.86-1.27 | 0.69 |
| Coronary artery operations | 0.74 | 0.69-0.79 | <0.001 | 0.98 | 0.92-1.05 | 0.64 | 1.23 | 0.98-1.53 | 0.070 |
| Diabetes | 1.05 | 1.01-1.09 | 0.021 | 1.25 | 1.20-1.30 | <0.001 | 1.56 | 1.38-1.75 | <0.001 |
| Hypertension | 1.09 | 1.06-1.12 | <0.001 | 1.24 | 1.21-1.28 | <0.001 | 1.33 | 1.21-1.46 | <0.001 |
| Hyperlipidaemia | 1.14 | 1.08-1.19 | <0.001 | 1.20 | 1.14-1.26 | <0.001 | 1.28 | 1.08-1.52 | 0.005 |

Stratified Cox proportional hazards regression, matched on age, sex, and GP practice (proxying deprivation). Time bands measured from individual condition diagnosis date. Reference group: individuals without the condition. HR, hazard ratio; CI, confidence interval.

### Sensitivity analyses

Sensitivity exclusions (cerebrovascular; neurological; severe mental illness) conditions across the study period removed 38,770 participants. Results were consistent with the primary analysis throughout. Similarly, when cases prior to 1 January 1987 were excluded (<1%, other than hypertension where n=17,241/9% of cases were excluded), this again made no difference to results.

## Discussion

### Overview

This study assessed the association between multiple cardiac and cardiometabolic conditions with subsequent dementia, including the first formal test of whether concurrent cardiac and cardiometabolic diagnoses confer synergistic risk. The main finding is that the greatest dementia risk is conferred by a first cardiovascular/cardiac diagnosis, with diminishing incremental risk from additional conditions. This pattern argues against a simple multi-hit model of cardiovascular contributions to dementia, and is consistent with substantially overlapping mechanistic pathways. The duration-dependent escalation of dementia risk due to CHD, hypertension, hyperlipidaemia, and most prominently diabetes, is a clinically important secondary finding. Effect sizes are generally more modest than those previously reported from recruited volunteer cohorts, highlighting the importance of routine data for accurate population- level risk estimation. We showed that apparently protective associations on dementia risk from certain conditions (e.g. atrial fibrillation, coronary artery operations, and/or heart failure) are likely due to competing risk i.e. where people die younger.

### Individual condition associations

Of the conditions meeting the proportional hazards assumption, the greatest risk was conferred by CHD which increased risk around 25%. This represents a smaller effect than a recent meta-analysis of prospective cohort studies which reported a combined OR of 1.55 (95% CI 1.20- 2.00) [19], though that combined sample was smaller than the CHD cohort alone in this study (N=34,270). The finding from this large, population-based sample may represent a more reliable estimate of population-level risk, and is closer to a recent UK Biobank study reporting an HR of 1.36 (95% CI 1.28-1.45) for all-cause dementia in people with CHD [20]. It is possible however that our nested case-control approach, studying people aged 60 years or over as of 2005, may bias associations towards healthier participants, and therefore associations towards null.

Hypertension was associated with a 17% increased risk consistent with the literature generally [8]. This is approximately in-keeping with a meta-analysis which reported a 21% dementia risk reduction associated with antihypertensive medication use [8]. Hyperlipidaemia was associated with a 17% increased risk, somewhat higher than a large meta-analysis (effect size 1.08, 95% CI 1.03-1.14) [21]. The evidence for MI is mixed [19]. The non-significant primary finding is consistent with prior studies reporting no significant effect [22], though survivor bias may contribute: MI survivors in an older adult cohort may represent a systematically healthier subgroup. The apparent protective associations for AF, CAO, and HF in overall models reflect competing risks from early mortality, confirmed by time-at-risk data and consistent with a prior competing risk analysis of AF [23].

### Greatest risk from a first cardiovascular diagnosis

Following grouping of conditions, individuals with any cardiac condition were at 13% increased risk of all-cause dementia and those with a cardiometabolic diagnosis at a 12% increased risk. People with both were at a 22% increased risk - greater than either individual category, but less than additive. The increased risk associated with multiple conditions is consistent with existing evidence that an increasing number of conditions is associated with greater cognitive decline or dementia risk [11–14]. However, prior multimorbidity subgroups have typically comprised around 100 participants [12]; where 82,473 participants in the current study had both a cardiac and a cardiometabolic condition, providing substantially more robust estimations.

The negative interaction indicates that the risk conferral effect of acquiring a second category of diagnosis, given an existing diagnosis in the other, is less than the effect of a first diagnosis from zero. The greatest dementia risk is therefore conferred by a first cardiovascular/cardiac diagnosis, with diminishing incremental risk from additional conditions. This pattern is most readily explained by substantial biological overlap in the pathways affected by cardiac and cardiometabolic conditions [6,9,16], where both condition types act on the same vascular, inflammatory, and/or metabolic pathways. The second category of disease therefore has less additional impact because those pathways are already affected. More intensive treatment and healthcare contact for patients with multimorbidity may further contribute to this dampening effect. Clinically, this argues for targeting cognitive risk reduction at the point of first cardiovascular/cardiac diagnosis, rather than waiting for patients to accumulate multiple conditions before flagging them as high-risk for dementia.

### Time-varying associations

Cox regression assumes that relative risk is constant over the follow-up period. In this study, this did not apply for diabetes, AF, CAO, and HF. Comparison of time at risk confirmed that death is a competing risk, particularly for AF, CAO, and HF - the conditions that appeared protective in outright initial models. Stratified regression found that AF, CAO, and HF were initially associated with decreased risk but showed directionally increased (though non- significant) risk at ≥15 years follow-up. This initial decrease likely reflects competing mortality, or protective factors such as anticoagulant treatment [5]; the diminishing protection over time suggests cumulative disease burden eventually outweighs these initial influences.

For diabetes, while associated with increased dementia risk across all follow-up periods, this varied substantially: from a 5% increased risk at 5-10 years to a 56% increased risk at ≥15 years. This is likely due to cumulative exposure to the vascular and metabolic complications of diabetes [7,24,25]. These findings are consistent with a UK Biobank study reporting a linear relationship between diabetes duration and dementia, with an HR of 2.15 (95% CI 1.60-2.90) for ≥15 years’ duration versus <5 years, independent of glycaemic control [24]. A German cohort study reported a U-shaped relationship with initial post-diagnosis risk reduction reaching its minimum at 4.75 years before increasing [25], potentially reflecting treatment effects or diagnostic delay until glycaemic control is established. The current findings, with a large diabetes cohort (N=61,008), add substantially to our understanding of duration-dependent dementia risk in diabetes.

### Strengths and limitations

Key strengths include the large, routine population-based sample; prospectively collected EHR data limiting recall and volunteer selection bias; incidence density matching controlling for age, sex, and deprivation by design; a long median follow-up (14.91 years); the exclusion of exposures within five years of dementia index; and systematic testing of the proportional hazards assumption with appropriate stratified modelling. The use of CPRD, with routine healthcare data broadly representative of the UK population [15], limits biases associated with volunteer datasets that can represent healthier, less diverse subsets. The total sample (N=347,436) is substantially larger than those used in other longitudinal multimorbidity and dementia research.

Several limitations warrant acknowledgement. First, residual confounding is likely: *APOE* genotype - the largest genetic risk factor for dementia after age [4] - was unavailable and likely underpins at least some of the cardiometabolic/dementia overlap. Lifestyle factors (smoking, physical inactivity, obesity) and medications (statins, antihypertensives, anticoagulants) could not be systematically adjusted for. However, *APOE* and lifestyle confounding would likely attenuate associations uniformly across exposure groups, so the relative comparison between conditions and the interaction estimate are probably less affected than absolute HRs. Second, IMD quintile was assigned at GP practice level: fewer than 45% of CPRD patients share the same IMD quintile as their practice [26], introducing potential error. Third, the sample skewed more affluent than the UK general population, likely reflecting survival selection - more deprived individuals with cardiovascular conditions may die before reaching dementia index age, or may be less likely to receive a diagnosis [27]. Fourth, without Hospital Episode Statistics linkage, secondary care dementia diagnoses may have been missed; combined with substantial underdiagnosis globally (around 62% in one meta-analysis [28]), this could result in underestimation of true associations. Our study was a case-control design which was enriched for dementia cases rather than following cardiovascular/cardiac patients prospectively; this may bias the association estimates. Finally, the inclusion of 2020 coincides with the COVID-19 pandemic, which disrupted primary care access and dementia diagnosis in older adults.

### Implications

The findings have important implications for public health policy makers and clinicians working in cardiovascular health. The Lancet Commission advocates action on modifiable risk factors that could prevent a significant proportion of dementia cases [4], and many - including smoking, physical inactivity, and excessive alcohol consumption - are shared with the cardiovascular conditions in this study. The finding that a first (cardiovascular/cardiac) diagnosis confers the greatest dementia risk reinforces the case for embedding cognitive assessment and targeted preventive action at the point of initial diagnosis, rather than reserving such action for patients with established multimorbidity and/or evidence cognitive decline in daily life.

Diabetes itself represents one of the modifiable risk factors on the Lancet Commission list [4]. The observed progression in dementia risk over the diabetes disease course is striking: type 2 diabetes prevalence has steadily increased in the UK in recent decades and significant under-diagnosis remains [29]. These results should strengthen public health efforts to prioritise early identification and intervention for type 2 diabetes, in part to support dementia prevention. Cognitive assessment could reasonably be considered an addition to the routine care processes for type 2 diabetes monitoring set out by NICE [30].

## Conclusion

This represents the largest population-based study to date of cardiovascular/cardiac multimorbidity and all-cause dementia risk, and the first to formally test whether concurrent cardiac and cardiometabolic diagnoses confer synergistic rather than additive risk in routine data. The central finding - that the greatest marginal dementia risk is conferred by a first cardiovascular/cardiac diagnosis, with diminishing incremental risk from additional conditions - argues against a cumulative, progressive model of cardiovascular neurodegeneration. Our findings are more consistent with substantially overlapping vascular, metabolic, and neuroinflammatory pathways. Duration-dependent escalation of dementia risk was observed across several conditions, most strikingly for diabetes but also for CHD, hypertension, and hyperlipidaemia, highlighting that cardiovascular disease duration - not merely presence - is a meaningful determinant of dementia risk. Apparently protective associations for atrial fibrillation, heart failure, and coronary artery operations were explained by competing mortality rather than ‘true’ biological protection. These findings strengthen the case for embedding cognitive assessment within cardiometabolic disease care from the point of first diagnosis, rather than reserving intervention for those with established multimorbidity. The progressive escalation of dementia risk with diabetes duration in particular supports prioritising early identification and sustained management of type 2 diabetes as a dementia prevention strategy.

## Author contributions

Conceptualization: Donald M. Lyall, Terry Quinn. Formal analysis: Thomas Donaldson, Jordan Canning, Donald M. Lyall. Methodology: Jordan Canning, Frederick K. Ho, Laura M. Lyall. Supervision: Donald M. Lyall. Writing - original draft: Thomas Donaldson, Donald M. Lyall. Writing - review & editing: Jordan Canning, Frederick K. Ho, Laura M. Lyall, Terry Quinn.

## Data Availability

Data cannot be shared publicly because they are provided by the Clinical Practice Research Datalink (CPRD) under a licence that does not permit the authors to share patient-level data. Data are available from CPRD (https://www.cprd.com) for researchers who meet the criteria for access to confidential data, subject to approval through the CPRD Research Data Governance process (this study: protocol 20_000212).

## Acknowledgments

The authors thank CPRD for access to the data used in this study, and the GP practices and patients whose anonymised records underpin this research. Thanks to Dr. Sharon Greenwood for facilitating aspects of this project.

## Competing interests

The authors have declared that no competing interests exist.

## AI statement

No AI was used in the conception, analysis or write-up of this manuscript.

## Data availability

Data cannot be shared publicly because they are provided by the Clinical Practice Research Datalink (CPRD) under a licence that does not permit the authors to share patient-level data. Data are available from CPRD (https://www.cprd.com) for researchers who meet the criteria for access to confidential data, subject to approval through the CPRD Research Data Governance process (this study: protocol 20_000212). The code lists used to define exposures, outcomes and sensitivity exclusions are provided in S1–S6 Tables.

## Abbreviations

AF: atrial fibrillation
CAO: coronary artery operation
CHD: coronary heart disease
CI: confidence interval
CPRD: Clinical Practice Research Datalink
EHR: electronic health record
GP: general practitioner
HF: heart failure
HR: hazard ratio
IMD: Index of Multiple Deprivation
MI: myocardial infarction
RERI: relative excess risk due to interaction

## Supporting information

**S1 Table.** CPRD GOLD exposure medcodes and descriptions. **S2 Table.** CPRD Aurum exposure medcodes and descriptions. **S3 Table.** CPRD GOLD outcome medcodes and descriptions. **S4 Table.** CPRD Aurum outcome medcodes and descriptions.

**S5 Table.** CPRD GOLD sensitivity exclusion medcodes and descriptions. **S6 Table.** CPRD Aurum sensitivity exclusion medcodes and descriptions. **S7 Table.** Cause-specific hazard ratios for dementia and death.

**S8 Table.** Multiplicative interaction between condition pairs for all-cause dementia.

**S9 Table.** Additive interaction (relative excess risk due to interaction) between condition pairs for all-cause dementia.

## Notes

### Competing Interest Statement

The authors have declared no competing interest.

### Author Declarations

CPRD has ethical approval from the Health Research Authority (Research Ethics Committee reference 05/MRE04/87) to provide anonymised patient data for observational research, so individual patient consent was not required. Access was approved through the CPRD Research Data Governance process (protocol 20_000212).

